# Feasibility of smartphone-based digital phenotyping to measure visual function and mental health outcomes in patients with inherited retinal diseases

**DOI:** 10.64898/2026.04.14.26350852

**Authors:** Lee Jones, Bethany Higgins, Kishan Devraj, David Crabb, Peter Thomas, Mariya Moosajee

**Author notes:** Corresponding author: Dr Lee Jones; Contact number; Address: 11-43 Bath St, London EC1V 9EL, United Kingdom. These authors contributed equally to this work (Lee Jones, Bethany Higgins).

## Abstract

This study evaluated the feasibility of collecting passive and active digital phenotyping data using the *OverSight* iOS application in individuals with inherited retinal diseases (IRDs), and explored associations between digital behavioural markers, visual function, and mental health. Participants with IRDs were recruited from Moorfields Eye Hospital (UK) and followed for 12 months. *OverSight* passively captures mobility data through HealthKit and typing-derived metrics through SensorKit. Participants completed patient-reported outcome measures (EQ-5D, NEI-VFQ-25, HADS, and MRDQ) within the app. Passive data included step count, walking speed, typing speed, total words typed, autocorrections, and sentiment word categories (anxiety, down, and health-related terms). Feasibility indices included enrolment, retention, and completeness of passive datastreams. Twenty-five participants were enrolled and 92% were retained at 12 months. Seventeen participants met the validity threshold for HealthKit data and 16 also met SensorKit thresholds. Median daily step count was 6,087, walking speed 1.18 m/s, and typing speed 2.19 characters/s. Age was negatively correlated with typing speed and anxiety-related word use, and photopic peripheral visual difficulty was negatively correlated with anxiety-and down-related word use. Digital phenotyping using *OverSight* was feasible over 12 months. Exploratory analysis suggest mobility, typing behaviour and sentiment markers may represent useful adjunctive indicators of functional vision and psychological outcomes in patients with IRDs.

## INTRODUCTION

Inherited retinal diseases (IRDs) are a heterogenous group of genetic eye disorders, affecting approximately 1 in 1000 people ^(1)^. Over 360 causative genes have been identified in the underlying pathology of IRDs which cause photoreceptor and retinal pigment epithelium dysfunction with associated vision loss. Visual impairment arising from IRDs can substantially affect everyday functioning, including mobility, daily activities, social participation, and psychological well-being. Retinitis pigmentosa (RP) is the most prevalent IRD, with an estimated global incidence of 1 in 3,000^(2)^. RP is characterised initially by rod photoreceptors degeneration leading to nyctalopia (night-blindness) and peripheral visual field loss, followed by cone involvement and eventual central vision impairment. Choroideremia (CHM) follows a similar pattern or peripheral-to-central deterioration and both conditions are marked by relatively preserved visual acuity (VA) until later disease stages ^(3–4)^.

Functional limitations are frequently reported by individuals with IRDs, particularly difficulties navigating in dim environments and avoiding peripheral hazards ^(5)^. Mobility is consistently identified as one of the most challenging domains in RP and a primary focus of low-vision rehabilitation ^(6)^. These challenges may contribute to reduced independence, lower vision-related quality of life, and elevated risk of anxiety and depression ^(7–9)^. Despite this burden, access to psychological support is inconsistent across services, and patients frequently report difficulty obtaining appropriate emotional or psychosocial care ^(10–15)^. This issue is particularly salient in conditions such as RP and CHM, where visual function and emotional states may interact ^(16)^. Taken together, the evidence suggests ophthalmology services, the busiest outpatient department in the National Health Service (NHS) ^(17)^, are not always equipped to identify and address the psychological dimensions of visual impairment.

In response to growing service demands, UK health policy emphasises digital transformation within the NHS to improve care delivery, enhance remote support, and promote timely access to clinical input ^(18, 19)^. One emerging innovation in medicine is the collection and analysis of behavioural and physiological data from personal digital devices, such as smartphones, referred to as digital phenotyping. Digital phenotyping incorporates both passive data (e.g., physical activity) and active patient self-reports (e.g., patient-reported outcome measures [PROMs]) to create a real-time, context-rich picture of a patient’s functional and mental health status. This approach has demonstrated utility in remote monitoring across psychiatric relapse prediction and prevention ^(20, 21)^, and post-treatment recovery in oncology ^(22, 23)^. In ophthalmology, digital phenotyping may offer ecologically valid insights into real-world functioning in domains such as mobility, which may serve as useful surrogate markers of visual function or overall health status ^(24)^. By capturing day-to-day behavioural patterns, digital phenotyping may also detect subtle changes not routinely identified during clinic-based assessments and thereby contribute to a more comprehensive understanding of patient experience.

This pilot feasibility study evaluated the use of a bespoke smartphone application (*OverSight*) designed to capture passive and active digital phenotyping data in individuals with IRDs ^(25)^. A secondary exploratory objective was to examine associations between digital behavioural metrics, self-reported visual functioning, and mental health indicators.

## METHODS

### Design

This pilot feasibility study involved onboarding individuals with RP and CHM to the *OverSight* smartphone app to assess enrolment, retention, and completeness of passive and active digital phenotyping datastreams. As this study was designed as a pilot feasibility evaluation, no formal sample size calculation was performed. The sample size was determined pragmatically based on the number of eligible patients available during the recruitment period. The study adhered to Strengthening the Reporting of Observational Studies in Epidemiology (STROBE) guidelines ^(26)^ (Supplementary Table 1), as recommended for feasibility studies ^(27)^. The protocol complied with the tenets of the Declaration of Helsinki and received approval from the North West–Greater Manchester South Research Ethics Committee (22/NW/0328).

### Recruitment

Participants were recruited from the Genetic Eye Disease Service at Moorfields Eye Hospital NHS Foundation Trust. Eligible participants were aged 16 years or older, had a molecularly confirmed diagnosis of RP or CHM, and a better eye VA of 6/60 Snellen (<1.0 logMAR) or better. Therefore, the better eye vision did not meet the UK threshold for severe sight impairment in the UK ^(28)^. This criterion was applied to minimise confounds related to severe vision loss in this initial feasibility evaluation. Opportunity sampling was used, with all eligible patients invited to participate. Demographic (age, gender, ethnicity) and clinical data (VA) were recorded at baseline by extracting measurements from the patient electronic medical record. Where Snellen VA was recorded, these values were converted to logMAR for reporting purposes.

### Procedure

Pre-screening was conducted by the clinical service lead (MM) to identify individuals meeting diagnostic and VA criteria. A brief secondary eligibility check confirmed (1) smartphone ownership, (2) use of an Apple device, and (3) installation of iOS 15 or later, ensuring compatibility with OverSight and reducing risk of operating system–related data capture limitations. Eligible individuals were provided with a participant information sheet outlining digital phenotyping procedures and smartphone datastreams collected. Written informed consent was obtained at the onboarding session, during which participants received an app tutorial. Onboarding was completed during an appointment or remotely once enrolment had been confirmed. Participants were not specifically instructed to open the application at regular intervals for passive data upload; instead, the app’s passive data capture was emphasised, with participants informed that routine smartphone activity data are collected unobtrusively ‘in the background’, and that active engagement would only be required to complete PROMs upon notification.

### *OverSight* application

*OverSight* was developed using Apple HealthKit and SensorKit frameworks. HealthKit provides aggregated physical activity and mobility data from the device, while SensorKit enables access to system-level keyboard metrics. *OverSight* was co-designed with individuals living with visual impairment, following user-centred design principles ^(29)^. Early end-user involvement informed accessibility, navigation, and notification features. Full development procedures are reported elsewhere^(25)^. Table 1 summarises sensor inputs, derived variables, and data types intentionally not collected due to privacy controls.

**Table 1.**
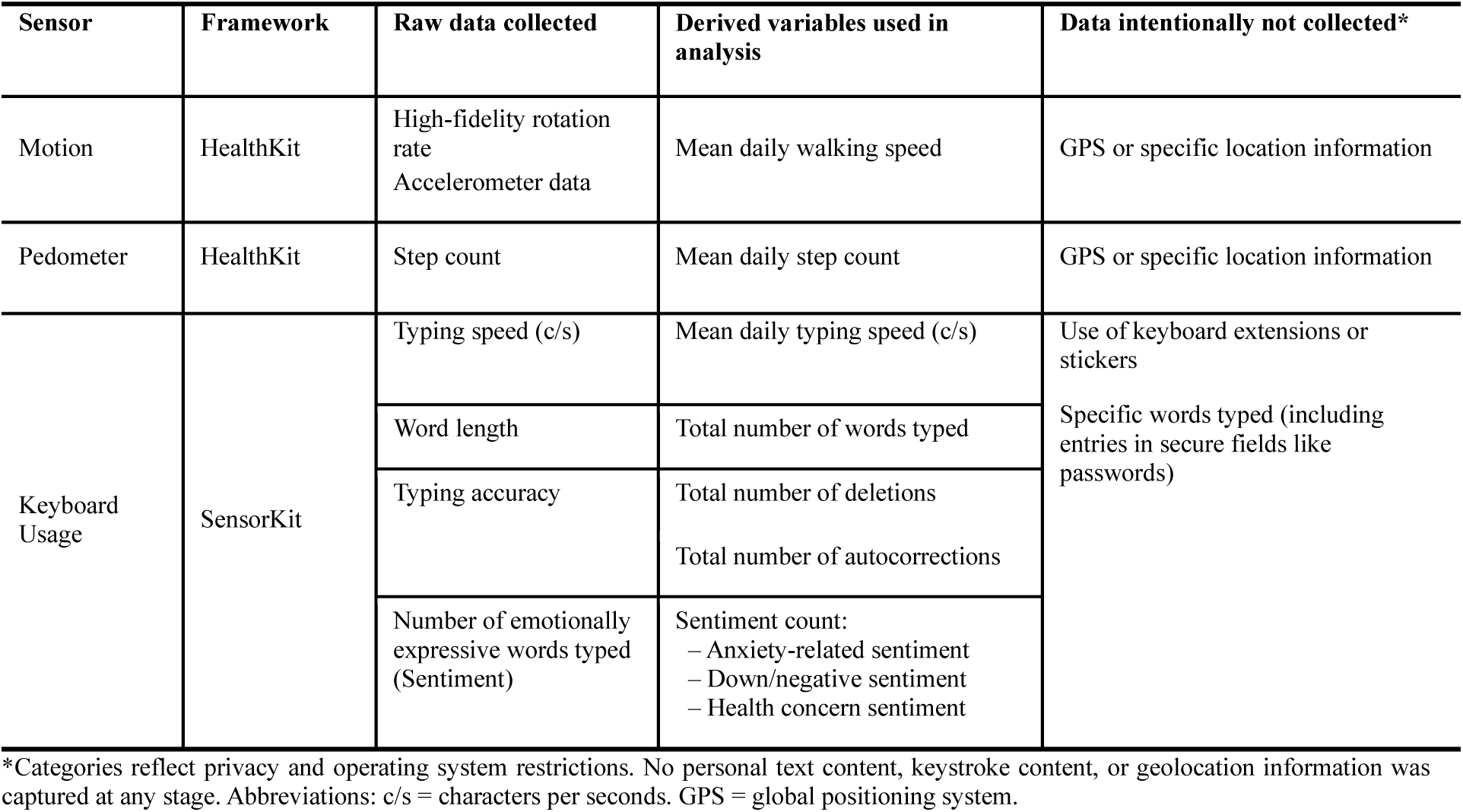
Sensor frameworks, data collected, and derived variables captured in the *OverSight* feasibility study.

### Data management

Passive datastreams from Apple HealthKit and SensorKit were collected over 12-months (March 2023–February 2024). PROM notifications were issued via the app at scheduled intervals. Data transfer to the encrypted University College London (UCL) server occurred only when the participant opened the app, initiating secure upload. Framework-specific handling was required due to differences in data retention. HealthKit data are stored locally on the device and were retrieved at study completion. SensorKit data, however, are retained for a maximum of seven days and automatically deleted thereafter unless transmission occurs. As a result, SensorKit capture was contingent on app opening within each seven-day window. Data quality procedures were implemented to address these operational differences and to ensure analytic consistency. For HealthKit mobility metrics, daily step and walking speed values were aggregated into weekly averages. A valid day was defined as ≥100 recorded steps, and a valid month as ≥25 valid days. Using these criteria, the most complete shared six-month interval (May–October 2023) was selected for analysis across participants. This interval was selected to maximise completeness and comparability across participants while avoiding the need for imputation, as data availability outside this shared period varied. SensorKit keyboard metrics were captured from discrete typing events and aggregated into daily values. A minimum of three days of extractable keyboard data was required for inclusion. Participants not meeting this threshold were excluded from keyboard-specific analyses.

### Outcomes

The primary outcome of this study was to determine the feasibility of collecting passive and active digital phenotyping data via the *OverSight* app. Feasibility was evaluated by examining the number of eligible participants who enrolled, the proportion retained over the 12-month study period, the completeness of passive (HealthKit, SensorKit) and active (PROM) data across the shared 6-month analytic interval, and the frequency of technical issues encountered during data collection. Retention was defined as participants remaining enrolled in the study and continuing to contribute either passive or active data at the 12-month follow-up. Thresholds used to define valid HealthKit and SensorKit data were applied separately for analytic purposes and reflect data completeness rather than retention. A secondary outcome was to explore the relevance of passive digital metrics in relation to participant characteristics and self-reported functioning. This included examining associations between passive datastreams and PROMs, as well as between passive metrics and demographic or clinical indicators, such as age and VA.

PROMs used in this study were the EuroQuol 5D (EQ-5D) health index ^(30)^, the National Eye Institute Visual Function Questionnaire (NEI-VFQ-25) ^(31)^, the Hospital Anxiety and Depression Scale (HADS)^(32)^ and the Michigan Retinal Degeneration Questionnaire (MRDQ) a validated measure designed specifically for individuals with IRDs ^(33)^. Standard scoring procedures were used for all PROMs. EQ-5D index scores range from values below 0 (health states worse than death) to 1 (full health), with higher scores indicating better health status. NEI-VFQ-25 composite scores range from 0 to 100, with higher scores indicating better vision-related quality of life. HADS anxiety and depression subscales each range from 0 to 21, with higher scores indicating greater symptom severity. MRDQ domain scores range from −3 to +3, with higher scores indicating greater perceived visual difficulty. No item-response theory or logit transformations were applied; summed scoring methods were used in accordance with instrument guidelines. Surveys were scheduled every 4-months for all measures (i.e., 3 timepoints) except the MRDQ, which was administered every 6-months due to its longer length (i.e., 2 timepoints). For analytic consistency, only the first completed survey for each measure was included in the present analysis. Most PROMs were completed within the shared six-month passive data interval, although a small number (n = 3) were completed outside this period.

### Statistical Analysis

To address the primary outcome of feasibility, we calculated the proportions of participants who enrolled and remained in the study at 12-months. Feasibility was further evaluated based on the number of participants meeting predefined data quality thresholds: at least six months of high-fidelity mobility data from HealthKit and a minimum of three days of keyboard data from SensorKit. For the secondary outcome, exploratory analyses examined associations between passive datastreams (step count, walking speed, typing speed, sentiment-related word use) and participant characteristics (age, better and worse eye VA), as well as PROMs (EQ-5D, NEI-VFQ-25, HADS, MRDQ). Spearman’s rank correlation coefficients were calculated to assess the strength and direction of associations. Multiple comparisons were corrected using the Benjamini–Hochberg procedure.

## RESULTS

### Overall feasibility

All 25 enrolled participants successfully downloaded the *OverSight* app and completed onboarding. Study retention over 12-month was 92% (23 out of 25), with attrition occurring only within the first 3 months and no subsequent withdrawals. Among these 23 participants, 74% (n = 17) met the HealthKit validity threshold (as defined in the methods). Smartphone operating system permission settings were the sole cause of incomplete HealthKit capture in the remaining six participants. Of the 17 participants with valid HealthKit data, 94% (n = 16) also met the SensorKit keyboard data threshold (≥3 days of extractable keyboard inputs). PROM completion was high overall, with 16 participants completing EQ-5D, NEI-VFQ-25, and HADS, and 13 participants (76%) completing all MRDQ domains. Feasibility outcomes are summarised in Figure 1.

**Figure 1.**
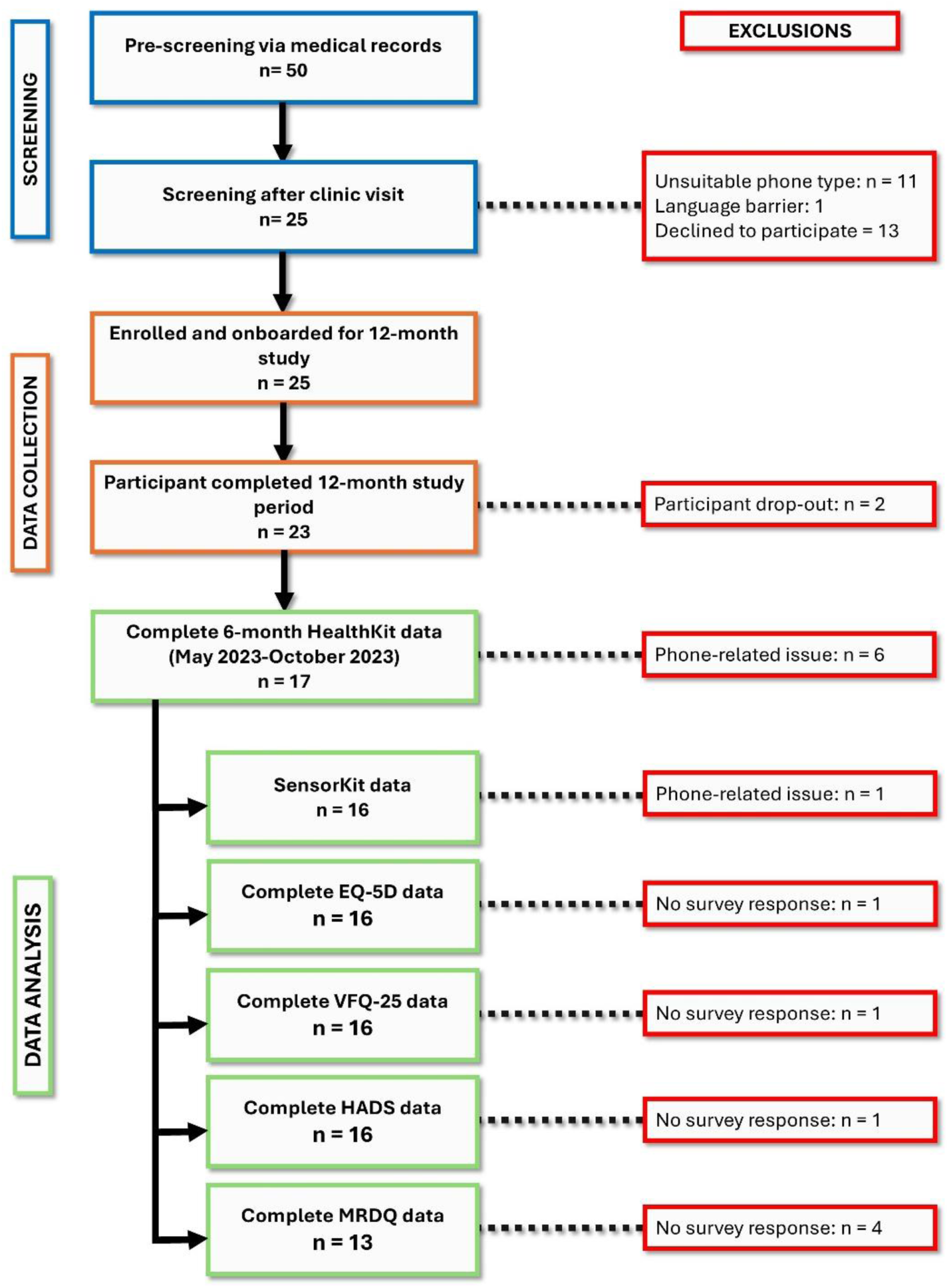
Participant flow and data completeness across the 12-month feasibility study: Of 50 individuals pre-screened, 25 were enrolled, with exclusions primarily due to incompatible smartphones or declining to participate. Retention at 12 months was 92% (n = 23). HealthKit data were analysable for 17 participants, with missingness attributable to device-related technical issues rather than participant withdrawal. Passive SensorKit data and survey measures (EQ-5D, VFQ-25, HADS) were complete for 16 participants, whereas MRDQ completion was lower (n = 13).

### Participant characteristics

Seventeen participants were included in the subsequent exploratory analysis. Demographic and clinical profiles are summarised in Table 2. The mean age was 39 years (SD ±16; range 16–65), with eight males (47%) and nine females (53%). All participants had an IRD, including RP (n = 14, 82%) and CHM (n = 3, 18%). Nine participants (53%) lived in urban settings, four (23%) in suburban environments, and four (23%) in rural areas. Employment status included nine (53%) in employment (one working from home), four (23%) retired, three (18%) unemployed, and one (6%) reporting other. In terms of visual function, mean better eye VA was 0.28 logMAR (SD ±0.19) and mean worse eye VA was 0.49 logMAR (SD ±0.46), indicating mild to moderate visual impairment among the sample.

**Table 2.** Participant demographics, clinical characteristics, residential setting and employment status.

| Study ID | Condition | Gender | Age bracket (years) | BEVA (logMAR) | WEVA (logMAR) | Ethnicity | Setting | Employment |
| --- | --- | --- | --- | --- | --- | --- | --- | --- |
| 01 | CHM | M | 16-24 | 0.1 | 0.1 | Caucasian | Suburban | Unemployed |
| 02 | RP | M | 35-44 | 0.3 | 0.6 | Caucasian | Urban | Employed, working from home |
| 03 | RP | F | 25-34 | 0.2 | 0.4 | Caucasian | Urban | Employed, not working from home |
| 04 | RP | M | 25-34 | 0 | 0.1 | Caucasian | Urban | Employed, not working from home |
| 05 | RP | F | 55-64 | 0.1 | 0.2 | Caucasian | Urban | Retired |
| 06 | RP | F | 45-54 | 0.3 | 0.7 | Caucasian | Suburban | Employed, not working from home |
| 07 | RP | M | 65-74 | 0 | 0.5 | Caucasian | Rural | Retired |
| 08 | RP | F | 16-24 | 0.3 | 0.5 | Caucasian | Urban | Other |
| 09 | RP | M | 55-64 | 0.2 | 0.3 | Caucasian | Rural | Retired |
| 10 | RP | M | 25-34 | 0.1 | 0.2 | Caucasian | Urban | Employed, not working from home |
| 11 | RP | F | 45-54 | 0.5 | 0.6 | Caucasian | Suburban | Employed, not working from home |
| 12 | CHM | M | 16-24 | 0.5 | 0.6 | Caucasian | Rural | Unemployed |
| 13 | RP | F | 45-54 | 0.6 | 1.7 | Caucasian | Urban | Employed, not working from home |
| 14 | RP | F | 35-44 | 0.4 | 0.4 | Caucasian | Urban | Employed, not working from home |
| 15 | RP | F | 55-64 | 0.4 | 0.5 | Caucasian | Suburban | Employed, not working from home |
| 16 | RP | F | 45-54 | 0.5 | 0.5 | Caucasian | Rural | Retired |
| 17 | CHM | M | 16-24 | 0.2 | 0.5 | Caucasian | Urban | Unemployed |

Abbreviations: BEVA = better eye visual acuity. WEVA = worse eye visual acuity. logMAR = logarithm of the Minimum Angle of Resolution. CHM = choroideremia. RP = retinitis pigmentosa.

### Passive data (HealthKit & SensorKit)

All 17 participants provided 6 months of valid mobility data (HealthKit), collected between May 2023 and October 2023. Validity criteria for monthly data completeness were applied as defined in the Methods; monthly data completeness ranged from 27 to 31 days. Median (IQR) weekly step count was 6,087 (3,891–8,344), and median (IQR) weekly walking speed was 1.18 m/s (1.08–1.26). Individual trajectories are shown in Figures 2–3.

**Figure 2.**
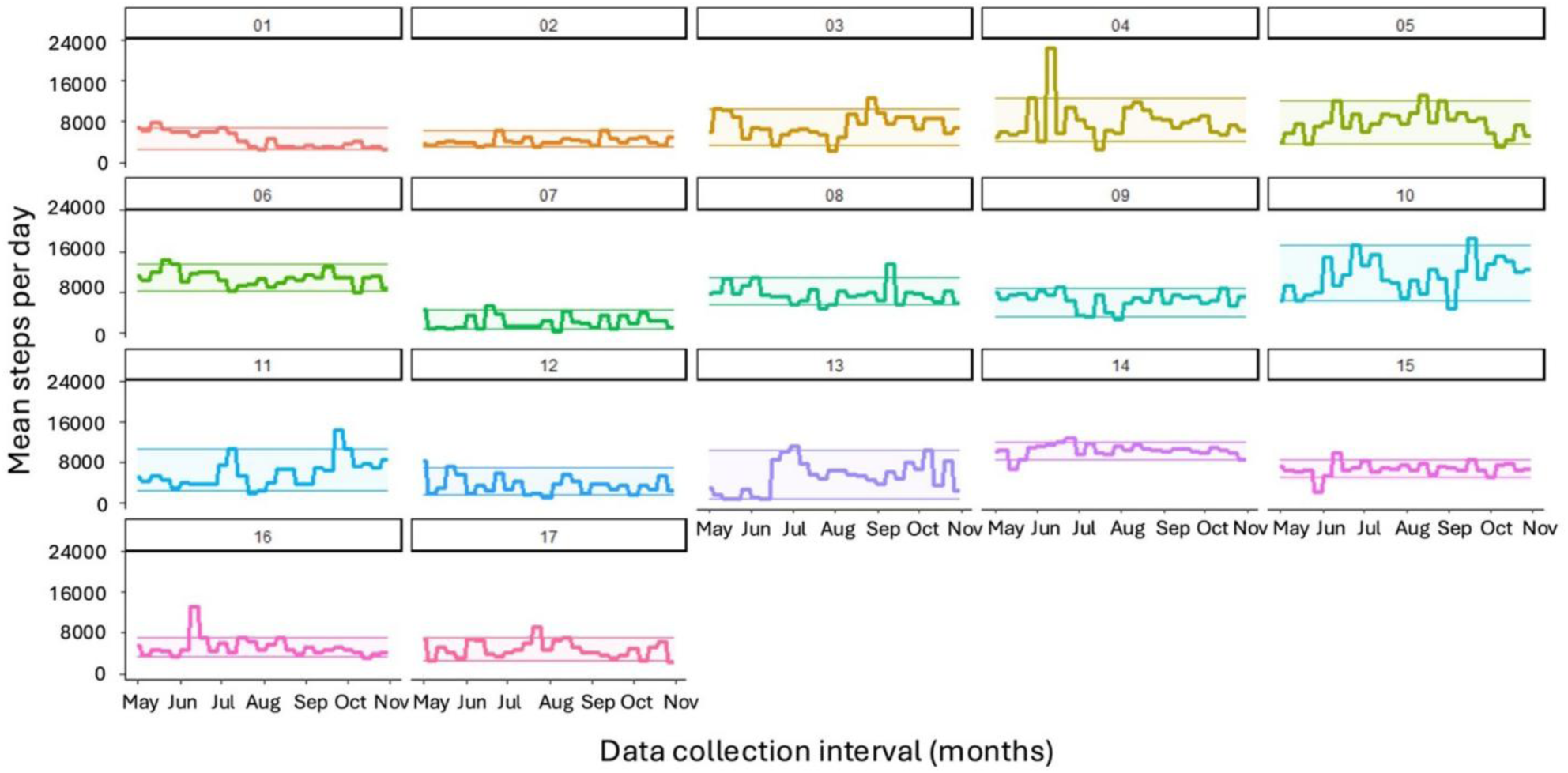
Six-month daily step count trajectories with individual variability: Mean daily step counts are displayed for each participant, with shaded bands marking personal 5th–95th percentile thresholds, indicating within-person fluctuation in physical activity.

**Figure 3.**
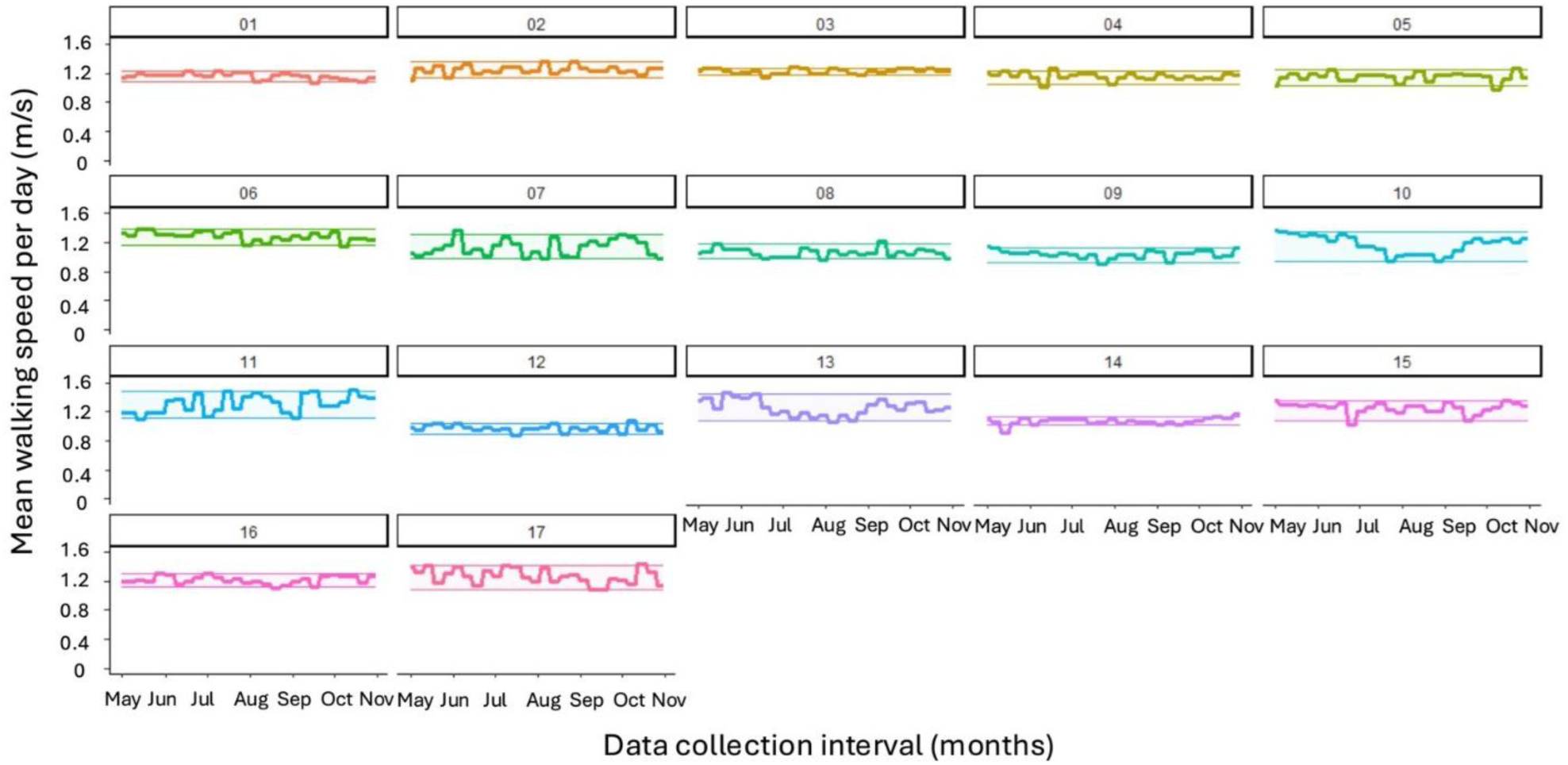
Six-month walking speed trajectories with individual variability: Mean daily walking speed (m/s) is displayed for each participant, with shaded bands marking personal 5th–95th percentile thresholds, indicating within-person fluctuation in mobility performance.

Sixteen of 17 participants provided valid keyboard-derived data (SensorKit). In total, 455 days of typing data were captured (median 38 days per participant; range 3–124). Median (IQR) typing speed was 2.19 characters per second (c/s) (1.88–2.98), and median (IQR) total daily typed words was 316 (159–855). Median (IQR) daily autocorrections were 51 (15–202). Sentiment word counts per day were fairly low overall: health-related words (median 1; IQR 0–4), anxiety-related words (median 0; IQR 0–1), and down-related words (median 1; IQR 0–3). Cohort-level distributions are shown in Table 3, and individual-level data in Table 4. Sentiment analysis was conducted using English-language lexical categories, and all participants were fluent English speakers.

**Table 3.** Cohort-level demographics, clinical measures, and passive and active digital phenotyping metrics.

|  | Median (IQR) | Range |
| --- | --- | --- |
| Age (years) | 41 (25, 51) | 16, 65 |
| BEVA (logMAR) | 0.3 (0.1, 0.4) | 0, 0.6 |
| WEVA (logMAR) | 0.5 (0.3, 0.6) | 0.1, 1.7 |
| Mean steps per day (weekly average) | 6087 (3891, 8344) | 304, 22292 |
| Mean walking speed per day (weekly average; m/s) | 1.18 (1.08, 1.26) | 0.88, 1.49 |
| Mean typing speed per day (c/s) | 2.19 (1.88, 2.98) | 1.07, 5.71 |
| Total words typed per day | 316 (159, 855) | 8, 5850 |
| Total autocorrections per day | 51 (15, 202) | 0, 1887 |
| Total health-related words typed per day | 1 (0, 4) | 0, 47 |
| Total anxiety-related words typed per day | 0 (0, 1) | 0, 24 |
| Total down-related words typed per day | 1 (0, 3) | 0, 26 |
| EQ5D health index score | 0.86 (0.81, 0.9) | 0.55, 1 |
| NEI-VFQ-25 composite score | 60.58 (41.64, 75.13) | 29.29, 90.08 |
| HADS – depression score | 4 (2.75, 5.25) | 1, 10 |
| HADS – anxiety score | 6.5 (4, 10) | 2, 16 |
| MRDQ – central vision | -0.16 (-0.53, 0.45) | -1.59, 1.12 |
| MRDQ – colour vision | 0.06 (-0.66, 0.96) | -1.35, 1.58 |
| MRDQ – contrast sensitivity | 0.12 (-0.48, 0.89) | -0.94, 1.70 |
| MRDQ – scotopic function | 0.88 (0.61, 1.73) | -1.29, 2.42 |
| MRDQ – photopic peripheral function | 0.81 (0.07, 1.13) | -1.26, 2.00 |
| MRDQ – mesopic peripheral function | 0.62 (0.39, 1.05) | -1.77, 2.42 |
| MRDQ – light sensitivity | -0.08 (-0.70, 0.80) | -1.82, 2.26 |

Abbreviations: m/s = meters per second. c/s = characters per second. BEVA = better eye visual acuity. WEVA = worse eye visual acuity. logMAR = logarithm of the Minimum Angle of Resolution. IQR = interquartile range. EQ5D = EuroQuol 5D. NEI-VFQ25 = National Eye Institute Visual Function Questionnaire. HADS = Hospital Anxiety and Depression Scale. MRDQ = Michigan Retinal Degeneration Questionnaire

**Table 4.** Participant-level mobility and keyboard-derived digital phenotyping metrics. Values represent within-person aggregated medians with interquartile ranges across the study period.

| Study ID | Mean steps per day (weekly average) | Mean walking speed per day (weekly average; m/s) | Mean typing speed per day (c/s) | Total words typed per day | Total auto-corrections per day | Total health-related words per day | Total anxiety-related words per day | Total down-related words per day |
| --- | --- | --- | --- | --- | --- | --- | --- | --- |
| Median (IQR) |  |  |  |  |  |  |  |  |
| 01 | 4065 (3056, 5975) | 1.17 (1.14,1.18) | 4.93 (4.74,5.05) | 1375 (542,2422) | 402 (164,734) | 2 (1,5) | 5 (1,7) | 6 (2,10) |
| 02 | 3899 (3743, 4678) | 1.23 (1.21,1.28) | 3.37 (3.29,3.43) | 924 (544,1335) | 235 (177,338) | 1 (0,1) | 3 (2,4) | 2 (1,5) |
| 03 | 6714 (5749, 9011) | 1.23 (1.20,1.26) | 4.82 (4.60,5.03) | 1841 (628,2681) | 248 (82,387) | 1 (1,6) | 5 (2,12) | 8 (5,50) |
| 04 | 6892 (5909, 9128) | 1.15 (1.13,1.19) | 4.39 (4.22,4.45) | 907 (825,1347) | 143 (133,207) | 1 (1,1) | 5 (4,6) | 5 (5,6) |
| 05 | 7618 (6086, 9429) | 1.16 (1.12,1.18) | 2.23 (2.14,2.33) | 293 (190,592) | 31.50 (23,66) | 0 (0,0) | 0 (0,1) | 2 (0,4) |
| 06 | 10833 (9914, 11617) | 1.29 (1.25,1.34) | 1.61 (1.53,1.71) | 132 (69,264) | 3 (1,5) | 0 (0,0) | 0 (0,1) | 0 (0,1) |
| 07 | 1743 (1024, 3442) | 1.10 (1.02,1.21) | 2 (1.92,2.13) | 210 (143,347) | 38 (22,62) | 0 (0,0) | 1 (0,1) | 0 (0,1) |
| 08 | 7510 (6263, 8117) | 1.06 (1.01,1.10) | 4.14 (4.11,4.41) | 1208 (1160,1424) | 231 (216,266) | 1 (1,3) | 7 (7,8) | 6 (3,7) |
| 09 | 6853 (5716, 7477) | 1.04 (1.01,1.07) | 3.02 (2.97,3.04) | 924 (794,1105) | 301 (265,378) | 0 (0,0) | 1 (0,3) | 1 (0,2) |
| 10 | 10597 (9178, 13420) | 1.20 (1.03,1.27) | — | — | — | — | — | — |
| 11 | 5269 (3868, 7299) | 1.33 (1.19,1.41) | 2.63 (2.54,2.73) | 1109.50 (439,1639) | 280 (120,474) | 2 (0,3) | 4 (2,9) | 6.50 (3,13) |
| 12 | 3456 (2032, 4208) | 0.98 (0.94,1) | 5.37 (5.14,5.60) | 1741 (563,2773) | 366 (149,572) | 3 (1,4) | 7 (2,9) | 8 (2,9) |
| 13 | 5381 (2594, 7865) | 1.25 (1.16,1.34) | 2.14 (2.08,2.16) | 1460 (1145,1797) | 337 (234,391) | 1 (0,1) | 0 (0,3) | 3 (1,5) |
| 14 | 10437 (9966, 11271) | 1.07 (1.03,1.09) | 2.96 (2.90,2.99) | 425 (121,943) | 69.50 (18.25,140) | 0 (0,0) | 1 (0,3) | 2 (0,4) |
| 15 | 6604 (6374, 7388) | 1.27 (1.21,1.29) | 2.16 (2.15,2.30) | 404 (402,493) | 69 (67,71) | 0 (0,0) | 1 (1,2) | 0 (0,1) |
| 16 | 4727 (4078, 5758) | 1.20 (1.18,1.26) | 2.92 (2.68,3) | 150.50 (20,411) | 18.50 (2,52) | 0 (0,0) | 0 (0,0) | 0 (0,2) |
| 17 | 4554 (3656, 5936) | 1.26 (1.18,1.34) | 4.77 (4.73,4.85) | 1348 (1153,3120) | 305 (274,704) | 1 (1,7) | 7 (3,17) | 8 (4,14) |

Abbreviations: BEVA = better eye visual acuity. WEVA = worse eye visual acuity. logMAR = logarithm of the Minimum Angle of Resolution. IQR = interquartile range. Note: Participant 10 did not contribute typing-derived data; values are missing for keyboard-based metrics.

### Active data (PROMs)

Across the 12-month study period, a total of 187 PROM completions were expected. Overall, 139 were completed (74%). Completion rates were 39/51 (76%) for EQ-5D, 38/51 (75%) for HADS, 38/51 (75%) for NEI-VFQ-25, and 24/34 (71%) for MRDQ. Sixteen participants completed baseline EQ-5D, NEI-VFQ-25, and HADS measures. Median (IQR) EQ-5D was 0.86 (0.81–0.90); NEI-VFQ-25 composite was 60.6 (41.6–75.1). Median (IQR) HADS Depression was 4.0 (2.75–5.25), and Anxiety was 6.5 (4.0–10.0). Based on HADS thresholds, depressive symptom severity ranged from normal to mild, while anxiety severity ranged from normal to severe. Thirteen participants (76%) completed the MRDQ. Domain medians (IQR) were: Central Vision −0.16 (−0.53–0.45), Colour Vision 0.06 (−0.66–0.96), Contrast Sensitivity 0.12 (−0.48–0.89), Scotopic Function 0.88 (0.61–1.73), Photopic Peripheral Function 0.81 (0.07–1.13), Mesopic Peripheral Function 0.62 (0.39–1.05), and Light Sensitivity −0.08 (−0.70–0.80). Individual PROM scores are shown in Table 5.

**Table 5.** Individual participant patient-reported outcome scores, including vision-specific and psychological measures. Values reflect within-participant score medians (IQR).

| Study ID | EQ-5D Index | NEI-VFQ-25 Score | HADS - Depression | HADS-Anxiety | MRDQ Domain Score |  |  |  |  |  |  |
| --- | --- | --- | --- | --- | --- | --- | --- | --- | --- | --- | --- |
|  |  |  |  |  | Central Vision | Color | Contrast | Scotopic Function | Photopic Peripheral Function | Mesopic Peripheral Function | Light Sensitivity |
|  |  |  |  |  | Median (IQR) |  |  |  |  |  |  |
| 01 | 0.90 | 84 | 2 | 11 | -0.93 | -0.02 | -0.06 | 1.08 | 0.03 | 0.53 | 0.81 |
| 02 | 0.81 | 64 | 10 | 7 | 0.26 | -0.46 | -0.26 | 0.58 | 0.93 | 0.56 | 0.04 |
| 03 | 1.00 | 81 | 2 | 4 | -0.34 | -0.08 | -0.94 | 0.83 | -0.30 | 0.67 | -0.80 |
| 04 | 0.94 | 66 | 2 | 5 | -1.38 | -0.72 | -0.55 | 0.41 | 0.18 | 0.21 | -1.82 |
| 05 | 0.78 | 45 | 6 | 10 | -0.07 | 0.55 | 0.29 | 0.70 | 0.72 | 0.48 | -0.33 |
| 06 | 0.89 | 57 | 4 | 9 | -0.24 | 0.14 | 0.60 | 1.77 | 1.28 | 1.09 | 0.59 |
| 07 | 0.87 | 54 | 5 | 4 | -0.25 | -0.84 | 0.38 | 0.93 | 0.83 | 0.74 | -0.21 |
| 08 | — | — | 1 | 6 | — | — | — | — | — | — | — |
| 09 | 0.85 | 29 | 4 | 6 | 0.51 | 1.05 | 1.11 | 2.42 | 1.86 | 2.42 | 1.75 |
| 10 | 0.81 | 70 | 5 | 7 | — | — | — | — | — | — | — |
| 11 | 0.69 | 40 | 7 | 16 | 0.76 | 1.37 | 0.98 | 1.89 | 0.93 | 0.93 | 1.00 |
| 12 | 1.00 | 90 | 3 | 2 | -0.59 | -1.35 | -0.86 | -1.29 | -1.26 | -1.77 | -0.83 |
| 13 | 0.55 | 38 | 3 | 3 | 0.53 | 1.58 | 1.22 | 1.63 | 1.19 | 1.51 | 0.75 |
| 14 | 0.86 | 42 | — | — | — | — | — | — | — | — | — |
| 15 | 0.86 | 73 | 3 | 4 | — | — | — | — | — | — | — |
| 16 | 0.76 | 31 | 9 | 14 | 1.12 | 1.02 | 1.70 | 2.10 | 2.00 | 2.13 | 2.26 |
| 17 | 0.92 | 80 | 4 | 10 | -1.58 | -0.72 | -0.55 | 0.43 | -0.36 | -0.01 | -1.59 |

Abbreviations: BEVA = better eye visual acuity. WEVA = worse eye visual acuity. logMAR = logarithm of the Minimum Angle of Resolution. IQR = interquartile range. EQ-5D = EuroQol 5-Dimensions Health Index. NEI-VFQ25 = National Eye Institute Visual Function Questionnaire. HADS = Hospital Anxiety and Depression Scale; MRDQ = Michigan Retinal Degeneration Questionnaire. HADS interpretation: Scores of 0–7 indicate normal symptom levels, 8–10 borderline (mild), 11–14 moderate, and 15–21 severe symptoms. Note: Participants 8, 10, 14, and 15 did not complete some PROMs; missing values are indicated by “—”

### Exploratory associations between digital phenotyping data and participant demographics

After false discovery rate correction, several associations remained. Age was negatively correlated with typing speed (ρ = –0.79, p-adj = 0.03), indicating that older participants tended to type more slowly. Age also correlated negatively with the average number of anxious-related words typed (ρ = –0.72, p-adj = 0.04), indicating greater use of anxious language among younger individuals. Photopic peripheral function (i.e., self-reported visual ability in the peripheral field under well-lit conditions) as measured by the MRDQ (higher scores indicating greater perceived difficulty), exhibited strong negative correlations with both the total number of anxious-related words and down-related words (ρ = –0.78 for both, p-adj = 0.04), indicating that participants reporting greater daytime peripheral vision difficulties used less negative emotional language when typing (see Figure 4).

**Figure 4.**
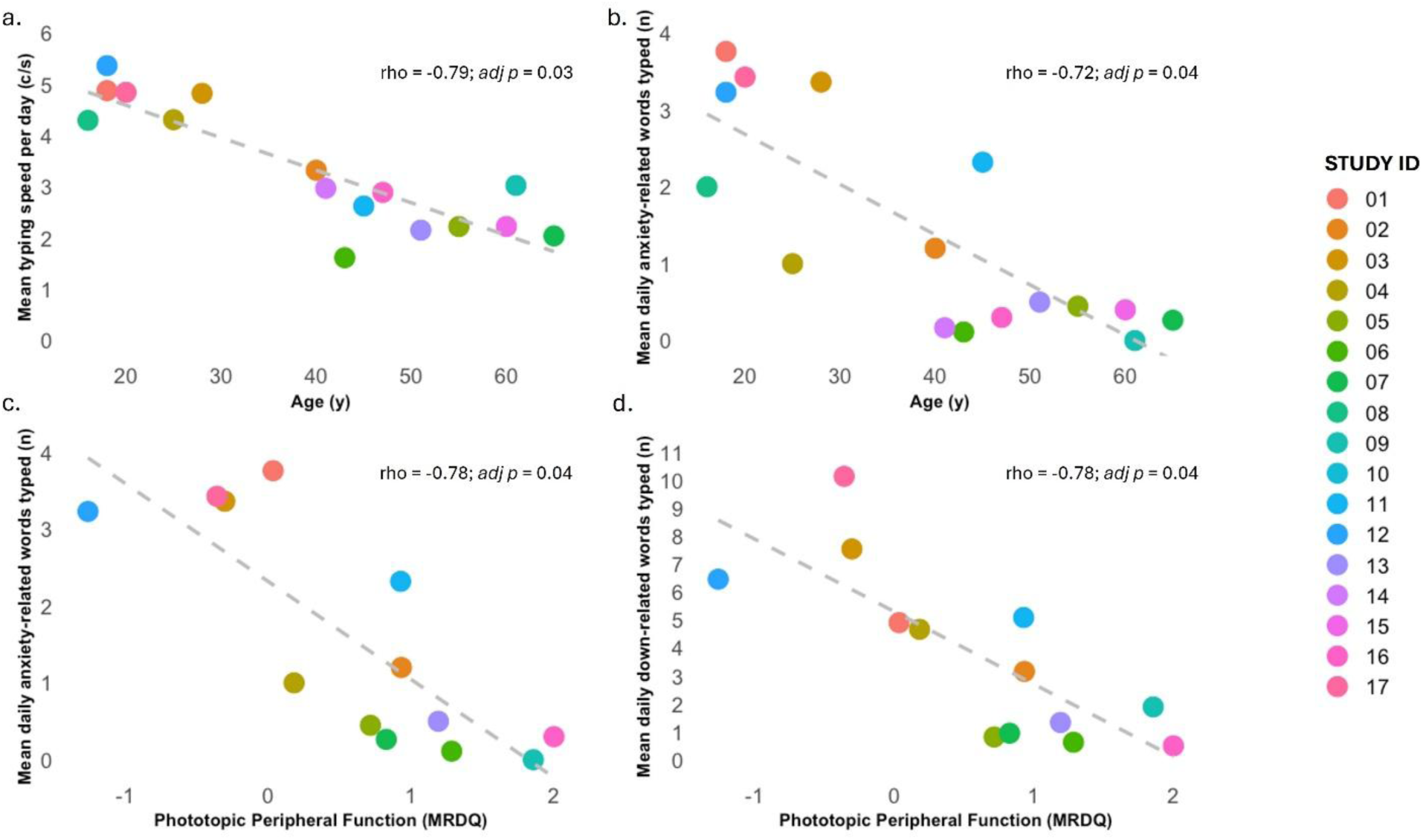
Significant associations between age, photopic peripheral function, and typing-derived behavioural markers: Scatterplots illustrate the significant Spearman correlations observed between: (a) age and mean typing speed, (b) age and mean daily anxiety-related words typed, (c) photopic peripheral function (MRDQ) and mean daily anxiety-related words typed, and (d) photopic peripheral function and mean daily down-related words typed. Each point represents an individual participant; dashed lines denote the fitted correlation trend. MRDQ scores range from −3 (indicating better photopic peripheral function) to +3 (indicating greater visual difficulty), with higher scores reflecting increased photopic peripheral impairment. All displayed relationships remained significant following multiple-comparison adjustment.

Several additional associations were present at trend level but did not meet corrected significance thresholds, including contrast sensitivity and typing speed (ρ = −0.69, p = 0.09), younger age and down-related word use (ρ = −0.70, p = 0.06), younger age and total words typed (ρ = −0.67, p = 0.07), and photopic peripheral visual function with total words typed, typing speed, and health-related word use (ρ = −0.69 for all, p = 0.08).

## DISCUSSION

This pilot study demonstrated the feasibility of using the *OverSight* smartphone app to collect passive and active digital phenotyping data in individuals with IRDs. Among participants retained at 12-months, sustained passive data capture meeting predefined HealthKit quality thresholds was achieved in 74% of participants, and 92% of these also met SensorKit thresholds. PROM completion was moderate to high, with 92% completing at least one PROM set (MRDQ: 76%). Collectively, these findings support the feasibility of multi-month passive and active behavioural data collection via *OverSight* in this population, while highlighting the importance of optimising longitudinal compliance and data completeness in future studies.

Mobility data indicated median weekly walking speed of 1.18 m/s and step count of 6,087 steps. While lower than typical step count and walking speed estimates reported in large general population datasets ^(34, 35)^, direct comparison was not the objective of this pilot. Given that smartphone-derived mobility data are influenced by environmental variables, seasonal variation, and local travel contexts, these findings primarily demonstrate that continuous mobility monitoring using *OverSight* is technically achievable in IRDs. Future controlled designs (e.g., matched cohorts) could more precisely assess mobility-related differences reported in IRD populations, including challenges with navigation, reduced confidence, and safety concerns ^(36)^.

Keyboard typing data collected via SensorKit revealed a median typing speed of 2.19 characters per second in this cohort. In comparison, Akpinar et al. ^(37)^ reported an average keystroke speed of 4.85 characters per second in a small but diverse sample of smartphone users (n = 48). The upper limit of typing speed in our cohort reached 5.71 characters per second, underscoring the relatively slower performance observed in individuals with IRDs. Typing speed varies by app context, with the fastest input typically seen in communication platforms and slower rates in productivity, social media, and utility applications ^(37)^. Performance is also shaped by environmental and cognitive factors, such as distractions, competing task demands, and whether the user is typing in their native language. Typing behaviour has been explored as a proxy for functional and psychological state, with smartphone-based keystroke features previously used to detect psychomotor change associated with depressive symptoms^(38)^. While cross-sectional comparisons between individuals are inherently variable, within-subject tracking of typing speed over time may provide more meaningful insight. Sustained declines or increased variability in typing behaviour could reflect changes in functional status, mood, or broader aspects of mental health, highlighting typing dynamics as a potentially relevant digital biomarker.

To address missingness and variability in the digital phenotyping data, predefined quality thresholds were applied to both HealthKit and SensorKit datastreams. While these thresholds improved analytic consistency, they also highlight a central challenge in digital phenotyping in achieving sustained, high-fidelity longitudinal capture. In this study, smartphone operating system permission settings were the primary determinant of incomplete passive data capture. Passive datastreams required users to grant and maintain access to device sensors, and in some cases permissions were not fully granted or access was interrupted when participants were logged out of the app and did not subsequently reopen it to re-enable permissions. Available keyboard data ranged from 3 to 124 days per participant, reflecting variability in engagement and technical constraints inherent to continuous passive collection. Missingness is well recognised in longitudinal health research ^(39)^, but its implications are heightened in digital phenotyping, where datastreams may inform individual-level clinical interpretation. Several strategies may help mitigate this issue in future studies, including structured onboarding procedures to verify permissions, clearer participant guidance, automated prompts when data streams are interrupted, and monitoring systems to identify data loss early so that participants can be supported in restoring access. Although statistical methods exist to account for missing data, their use in digital phenotyping remains inconsistent, and the consequences for clinical validity are not yet fully established ^(40)^. Standardised approaches to managing missingness will therefore be essential for future work aiming to advance digital phenotyping toward clinical applicability.

Our analysis identified associations between emotionally relevant language used during keyboard typing, age, and self-reported visual functioning. Participants reporting greater perceived difficulty in photopic peripheral vision on the MRDQ showed significantly lower daily use of anxiety-and low mood–related terms, and those with greater peripheral and contrast sensitivity difficulties tended to type fewer words and at a slower speed. This pattern may reflects reduced expressive output rather than reduced emotional distress, given that individuals with poorer visual function typed less overall. In contrast, younger participants demonstrated significantly greater use of anxiety-related terminology and a trend toward increased down-related language, indicating higher emotional expressivity in typed communication. While exploratory, these findings suggest that sentiment tracking via *OverSight* may offer insight into the lived emotional experience of individuals with IRDs. This is particularly relevant in the UK context, where rising demand on mental health services limits timely access to support ^(41)^, and extended waiting times are associated with poorer outcomes, including reduced treatment response and increased risk of comorbidities ^(42)^. Digital approaches therefore have potential to complement existing care pathways. For example, smartphone-based systems such as EmoKey have demonstrated the capacity to classify emotional states (e.g., happiness, sadness, stress, relaxation) using machine learning methods with accuracy of up to 78% ^(43)^, highlighting emerging possibilities for digital mood monitoring.

These findings should, however, be interpreted cautiously. Sentiment word counts were low overall, and MRDQ data were available for only a subset of participants, limiting statistical power. Emotionally expressive language captured through keyboard input cannot be assumed to reflect clinical need, as some individuals who express higher levels of negative sentiment may not require intervention, while others who type minimally may nonetheless experience significant psychological distress. Passive sentiment monitoring may therefore be best considered as an adjunctive indicator rather than a standalone measure. Such approaches could be most relevant in specific clinical contexts such as postoperative recovery, vision rehabilitation programmes, or periods of documented functional decline, where early recognition of distress may improve patient support. There is also increasing interest in integrating patient-centred outcomes into ophthalmic clinical trials; sentiment-derived metrics may complement conventional functional endpoints and contribute to more holistic assessment of treatment impact ^(44)^.

Age-related differences in typing speed observed in our sample provide preliminary support for the sensitivity of digital phenotyping in capturing real-world behavioural variation. This finding aligns with evidence that healthy ageing is associated with psychomotor slowing ^(45, 46)^, and therefore offers a useful validity check for the passive datastreams collected via *OverSight*. In contrast, no significant associations were observed between passive metrics and VA. This may in part reflect the small sample size and limited statistical power of this feasibility study, but is also consistent with longstanding recognition that acuity alone is a limited indicator of functional visual experience, compared to measures such as visual fields. More precise disease-staging methods, such as high-resolution clinical imaging, including spectral-domain optical coherence tomography and fundus autofluorescence, offer enhanced structural characterisation of IRDs and should be incorporated in future work. Because between-person comparisons are influenced by routine variation and lifestyle differences, longitudinal within-subject designs are likely to be more informative. Pairing digital phenotyping with staged imaging over extended follow-up may therefore provide a more sensitive means of identifying clinically meaningful functional change as disease progresses.

PROM completion was moderate to high, with most participants completing at least one scheduled PROM set and an overall completion rate of 74% across repeated assessments. This level of engagement is consistent with existing mHealth research showing that smartphone platforms can support large-scale collection of patient-reported health information ^(47)^. The literature highlights accessibility and usability as key determinants of PROM completion and data quality ^(48)^, and *OverSight* was co-designed with individuals living with visual impairment to ensure that interface features aligned with user needs. These results are consistent with prior evidence demonstrating that accessible, intuitively designed digital tools can promote adherence among people with visual impairment ^(49)^. Inclusive design, early user involvement, and the strong commitment shown by participants likely contributed to the high engagement observed in this study. At the same time, dropout rates in mHealth studies tend to be higher in routine deployment than in controlled study environments ^(50)^, and therefore scalability requires further evaluation. Encouragingly, our previous work has shown that individuals with IRDs and their families view the concept of digital phenotyping as acceptable ^(51)^. Completion of the MRDQ was lower than for the other PROMs, which may reflect the greater length of this instrument and the potential for survey fatigue in longitudinal studies. Questionnaire burden is an important consideration in digital phenotyping research, where repeated measures are often required, and future studies may benefit from optimising survey frequency or using shorter instruments to support sustained engagement.

PROMs were administered at intervals of 4–6 months in this feasibility study, which may not capture short-term fluctuations in psychological symptoms. More frequent assessment approaches, such as ecological momentary assessment (EMA), have been shown to improve sensitivity to variation in mood and may represent a more appropriate strategy for monitoring mental health in future digital phenotyping studies ^(52)^. Clinical relevance in mental health often depends not only on the presence of symptoms but also on their persistence over time, and more regular assessment may therefore be important for detecting sustained psychological symptoms. Determining optimal assessment frequency while minimising participant burden will be an important consideration in subsequent research phases.

*OverSight* aligns with ongoing developments in ophthalmology aimed at expanding remote monitoring and improving access to care. Over the past decade, remote assessment of eye diseases has advanced through technologies enabling evaluation outside traditional clinical settings ^(53–56)^. The widespread availability of smartphones has facilitated apps capable of remotely assessing visual function, including VA ^(57, 58)^. Digital phenotyping extends this landscape by capturing behavioural and physiological data that may offer complementary insights into visual functioning and broader aspects of health and well-being. While this study focused on smartphone-derived mobility and typing-based metrics, future work may incorporate additional datastreams relevant to visual functioning and explore integration with other sensors, such as wearables.

With respect to limitations, the small sample size, typical of pilot work, limits generalisability. The sample was restricted to individuals with a VA better than 1.0 logMAR in the better-seeing eye, which ensured adequate app usability but does not reflect the full spectrum of IRDs, particularly more advanced visual loss. Recruitment from a single specialist centre may also have introduced response bias, as participants’ established clinical relationships could have positively influenced engagement. Variability in smartphone use, device type, and interaction behaviours likely contributed to noise within passive datastreams. Although baseline questioning confirmed that participants carried their phones most of the time, we did not assess the specific contexts in which devices were worn or positioned (e.g., on-person vs nearby), which would improve interpretability but warrants testing in a larger trial. Passive data completeness was partly influenced by smartphone operating system permission settings, as participants could modify sensor access during the study or experience interruptions in access (e.g., after logging out of the app). While this reflects real-world conditions in digital phenotyping research, it contributed to variability in data capture and should be considered when interpreting feasibility outcomes. Passive data upload depended on participants periodically opening the app, particularly for SensorKit data with limited retention windows. Participants were not routinely prompted to open the app outside scheduled self-report assessments, allowing feasibility to be evaluated under relatively naturalistic conditions. However, this reliance on active user behaviour may limit scalability without further automation. Future iterations may benefit from optimised scheduling of low-burden self-report measures, such as EMAs, which could both improve the sensitivity of psychological measures while also promoting more regular app engagement. Increasing digitalisation of healthcare and growing familiarity with patient-generated health data may further support the feasibility of such approaches in future. Although participants were enrolled for 12 months, analyses focused on the six-month interval with highest data completeness to ensure comparability. This highlights the importance of considering optimal monitoring intervals in digital phenotyping, which remain to be established and are likely to depend on the clinical context and the outcomes of interest. Selection of a shared six-month analytic interval improved comparability across participants but limits conclusions about longer-term longitudinal patterns. Decisions regarding preprocessing, including definitions of valid days and valid months, are likely to influence both analytic outcomes and the feasibility of scaling digital phenotyping approaches in clinical settings. Future studies with larger samples and improved automation of data capture will be better positioned to evaluate continuous long-term monitoring. Although most PROMs were completed within the passive data interval, minor variation in the timing of active and passive data collection may have influenced observed associations. Sentiment analysis was based on English-language lexical categories, and findings may not generalise to multilingual populations or to individuals who primarily communicate in other languages. This should be considered in future research where multilingual populations are recruited. Lastly, the *OverSight* app was compatible only with Apple iOS devices, which may introduce digital exclusion and limit generalisability. While a single-platform approach was appropriate for this early feasibility evaluation to ensure controlled testing, future development across multiple operating systems will be important to support equitable access and wider clinical implementation.

This pilot study demonstrates the feasibility of using the *OverSight* app to collect passive and active digital phenotyping data in individuals with IRDs. Data acquisition via Apple’s iOS frameworks was technically achievable and supported by high participant compliance and acceptable overall completeness. These findings indicate that smartphone-based collection of mobility, typing, and patient-reported outcomes is practicable in this population. As feasibility outcomes, the results should be interpreted as preliminary. Larger, methodologically powered studies are required to assess generalisability, optimise data capture, and determine the clinical utility of digital phenotyping for remote monitoring in ophthalmology.

## DECLARATION STATEMENTS DATA AVAILABILITY

The datasets generated and/or analysed during the current study are not publicly available due to the inclusion of identifiable participant information and restrictions imposed by the study’s ethical approval. De-identified data may be available from the corresponding author upon reasonable request and subject to institutional data sharing agreements.

## CODE AVAILABILITY

The code for this study is not publicly available but may be made available to qualified researchers on reasonable request from the corresponding author.

## Data Availability

De-identified data may be available from the corresponding author upon reasonable request and subject to institutional data sharing agreements.

## ACKNOWLEDGEMENTS

This study was funded by UKRI Economic and Social Research Council (grant number ES/W006510/1), Wellcome Trust (grant number 205174/Z//16/Z), Thomas Pocklington Trust, and the NIHR Moorfields Biomedical Research Centre. The funder played no role in study design, data collection, analysis and interpretation of data, or the writing of this manuscript.

## AUTHOR CONTRIBUTIONS

L.J. collected and analysed the data, drafted the manuscript, secured funding, and conceptualised the study. B.H. conducted data preparation, visualisations and analysis, and drafted the manuscript. L.J. and B.H. contributed equally to this work. K.D. collected the data. D.P.C edited and reviewed the manuscript. P.B.M.T. edited and reviewed the manuscript. M.M. conceptualised the study, edited and reviewed the manuscript. All authors have read and approved the final version of the manuscript.

